# Evaluation of the Hospital Readmissions Reduction Program

**DOI:** 10.1101/19011734

**Authors:** Rohan Khera, Yongfei Wang, Susannah M. Bernheim, Zhenqiu Lin, Sharon-Lise T. Normand, Harlan M. Krumholz

**Author notes:** **Corresponding Author** Dr. Harlan M. Krumholz; 1 Church St., Suite 200, New Haven, CT 06510.

## Abstract

**Background:** There is conflicting evidence about whether the Hospital Readmission Reduction Program (HRRP) is associated with an increase in mortality.

**Methods:** In a cohort of Medicare beneficiaries hospitalized with heart failure (HF), we compared two published approaches to evaluating the association of HRRP and risk-adjusted 30-day mortality, including changes in average mortality across periods and changes in slope of monthly mortality rates across discrete periods. We also tested various methods with simulated data that was designed with an inflection in mortality.

**Results:** We identified 4,313,523 hospitalizations for HF, 1,788,219 for AMI, and 3,758,111 for pneumonia. Monthly slope-change models identified an increase in mortality for HF and pneumonia in the pre-HRRP period (P<.001 for slope-change). The changes in average mortality across the four time periods model found an increase in mortality for HF and pneumonia in the HRRP anticipation period (post-announcement, pre-implementation) as well as following HRRP implementation. However, under those conditions, our simulations reveal that method errs in identifying the timing of a change. Varying the data sources and risk-adjustment strategies had no significant effect on the results.

**Conclusion:** A national policy incentivizing efforts to reduce readmission did not increase the risk of mortality.

## BACKGROUND

Recent studies evaluating the Hospital Readmission Reduction Program (HRRP), a federal policy that incentivizes reducing hospital readmissions, have reported conflicting results about its association with patient mortality after discharge for patients with heart failure and pneumonia.^1-11^ Studies are consistent in reporting that HRRP is not associated with an increase in mortality for patients with acute myocardial infarction.

Amid the uncertainty, some experts are calling for the HRRP to be revoked. Others, including Medicare Payment Advisory Committee (MedPAC) – a bipartisan committee that advises the government on Medicare – concluded that the policy has averted many readmissions, saved billions, and has not caused harm.^1,12^ There is a need to investigate the source of conflicting results and to determine if there are methodological issues that explain the differences. A few studies have evaluated the association of HRRP and mortality,^2,4,5,8,9^ but only two prominent published studies have used the entire complement of discharges from the Centers for Medicare and Medicaid Services (CMS) and sought to pinpoint changes in temporal trends in mortality in relation to the HRRP announcement and implementation. One study evaluated monthly changes in the slope of mortality rates over time,^3^ and another compared differences in average mortality rates between periods before the announcement of HRRP, during the announcement, and then implementation.^5^

There are several methodological differences between studies that may explain the different conclusions. One influential study, which inferred that there was an increase in mortality associated with HRRP,^5^ compared differences in average mortality rates between periods before the announcement of HRRP, during the announcement, and then implementation.^5^ In contrast, another study, which failed to find such an effect, evaluated monthly changes in the slope of mortality rates over time.^3^ A central assumption of the latter method is that if there are no temporal changes within those periods that could lead to an inability to identify accurately the time of the change.^13^

Accordingly, we conducted a series of data experiments to test the effect of the analytic choices on the temporal trends in patient risk and mortality in the period spanning the announcement and implementation of the HRRP. We additionally tested the assumption that there was no temporal trend within the periods, which is essential to the period-wide assessment method. We developed simulated data in which we created an inflection in mortality and tested the ability of the two methods to correctly identify the inflection and its timing. We also investigated whether results were sensitive to the choice of source data and strategies to handle changes in risk factors as claim reporting standards changed concurrently with HRRP’s announcement.^14,15^

## METHODS

### Data Source and Study Population

We used the Medicare Standard Analytic Files including data from April 2005 through March 2015 to capture periods included in recent studies evaluating the HRRP. We included hospitalizations among fee-for-service Medicare beneficiaries, ≥65 years of age, with a principal discharge diagnosis of HF, AMI, or pneumonia – 3 conditions included in the HRRP. The study population was consistent with the population under the purview of the program.^3,16^

### Study Periods

We divided the 120-month period between April 2005 and March 2015 into four 30-month periods based on the discharge date of the inpatient admissions: (a) baseline (April 2005-September 2007); (b) pre-HRRP (October 2007-March 2010), a 30-month period before HRRP announcement; (c) HRRP anticipation (April 2010-September 2012), a 30-month period between HRRP’s announcement and implementation; and (d) HRRP penalty (October 2012-March 2015), when the Medicare penalized hospitals for higher than expected readmissions rates. These consistent periods of observation allowed us to evaluate for changes in outcomes before the introduction of HRRP, and after its announcement and subsequent implementation.^5^

### Overview of Data Experiments

We hypothesized that differences in results may derive from differences in study design, data source, or the approach to inpatient codes for risk adjustment. We constructed data experiments to test these 3 potential mechanisms (**Figure 1**). To permit head-to-head assessments, we harmonized the data, keeping the study population and period of observation consistent across experiments. We examined the same outcome – all-cause post-discharge 30-day mortality – across all experiments. Finally, the experiments were conducted in parallel, such that we isolated the effect tested in one experiment to the other two.

**Figure 1:**
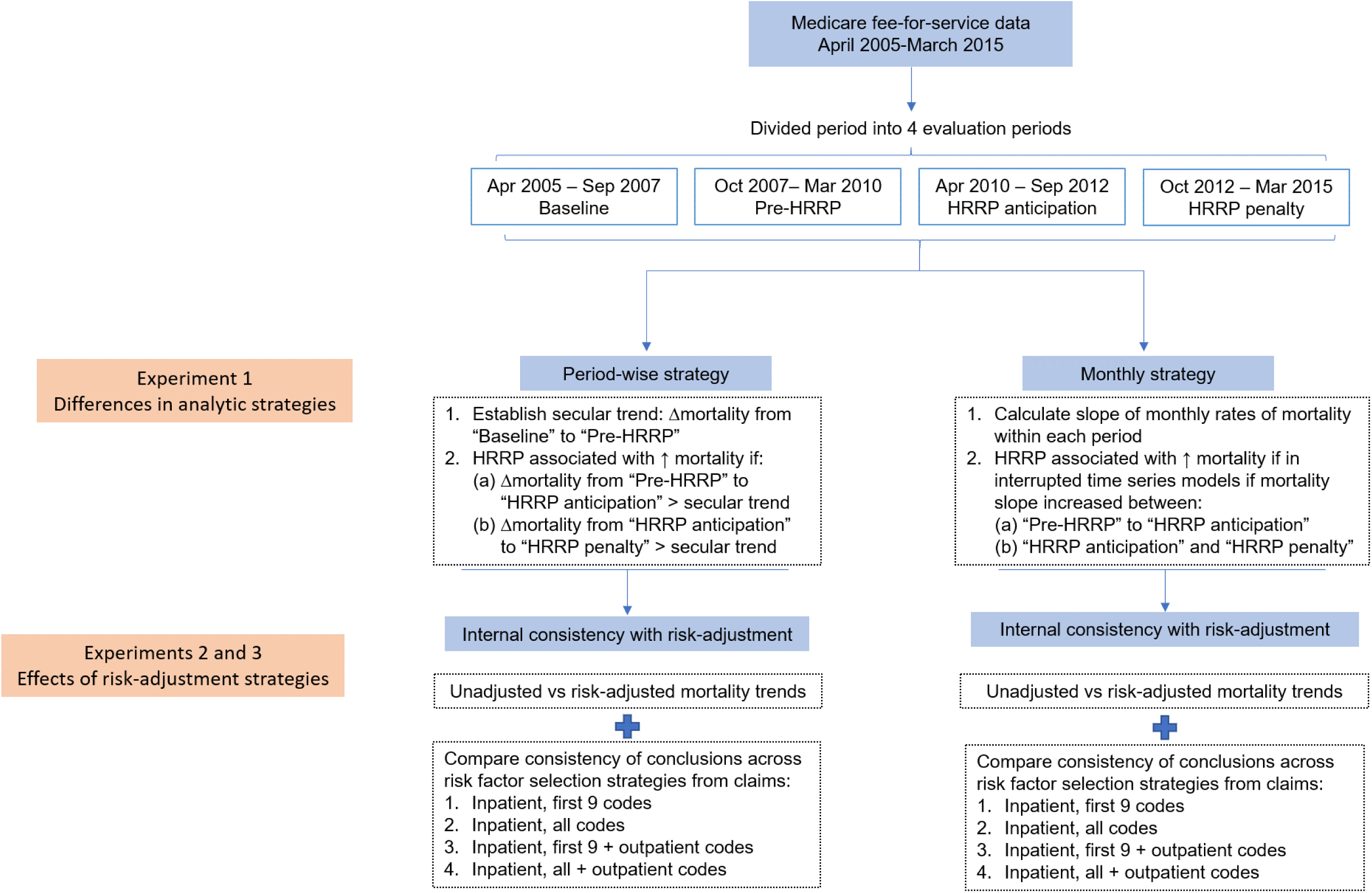
Overview of Data Experiments.

#### Experiment 1: Mortality patterns in period-averaged vs monthly assessments

To test whether differences in study design produced different results, we adopted a pre-post design that evaluated whether average risk-adjusted mortality differed among the 4 study periods ignoring within-period temporal trends as previously reported.^5^Using the same 4 periods, we evaluated whether risk-adjusted monthly mortality differed within the study periods, thus explicitly modeling within-period time trends while evaluating changes across periods in an interrupted time series framework (**Figure 1**). We determined whether the two approaches produced different results using the same data, the same study periods, and the same approach of risk-adjusting data, thereby isolating the effects of the modeling strategy. We also tested whether the assumption about absence of within-period trends, which is necessary for studying the effects of HRRP on average mortality, were satisfied.

#### Experiment 2: Risk-adjustment with Inpatient vs Inpatient and Outpatient Claims

We identified comorbidities used in the CMS risk-adjustment models from 3 sources: (1) secondary diagnoses in the index hospitalization, (2) all diagnoses and procedures during hospitalizations over the preceding 12 months, and (3) diagnosis and procedure codes across all outpatient and professional claims over these 12 months. One study that found an increase in mortality with HRRP, only used inpatient data (sources 1 and 2 above) to assess patient risk, compared with another that used all inpatient and outpatient data (all 3 sources) and did not find an increase in mortality with HRRP. We assessed whether the differences in the association with mortality arose due to different data sources for risk-adjustment.

#### Experiment 3: Changes in Number of Codes on Inpatient Claims

We tested the effect of how studies handled changes in CMS data transaction standards around HRRP announcement, when the number of secondary diagnosis codes and procedure codes on inpatient claims increased from 9 to 24, and 6 to 25, respectively. This experiment was an extension of Experiment 2, wherein the comparisons between inpatient-only vs inpatient and outpatient strategies to identify risk factors were stratified into two subgroups each, based on the use of 1) the first 9 secondary diagnosis codes and first 6 procedure codes to derive risk factors even when a larger number of codes were available; and 2) using all the inpatient codes that were available.

### Statistical Analysis

We described the mean monthly number of risk factors and mean mortality risk across the 4 periods. The risk scores were based on regression coefficients derived from two logistic regression models with post-discharge 30-day mortality as outcome – constructed in parallel using covariates from inpatient data vs inpatient and outpatient data. The monthly risk score represented the sum of risk factors weighted by their contribution to mortality averaged across hospitalizations for each month.

For the period-wise analyses that ignored within-period monthly trend approach in Experiment 1, we calculated the average difference in mortality across all months between the baseline (period 1) and pre-HRRP (period 2), pre-HRRP and HRRP anticipation (period 3), and HRRP anticipation and penalty (period 4) periods. The change between periods 1 and 2 was assumed to represent the secular trend for mortality, and the excess mortality changes between periods 2 and 3 and periods 3 and 4 were assumed to be temporal changes in mortality with HRRP announcement and HRRP penalties. The differences in changes were assessed as marginal effects in regression models that modeled period as the covariate of interest, and mortality as the outcome (**Supplementary Appendix)**.

In the second approach, we used a slope-change interrupted time series framework to evaluate changes in unadjusted and risk-adjusted mortality across transitions between the 4 periods defined in the first approach.^17^ We constructed a linear spline using the monthly post-discharge mortality rates within each of 4 periods outlined above, with knots at the transitions between these periods. We evaluated for changes in slope of mortality at the knots between these periods, including between the baseline and the pre-HRRP periods, the pre-HRRP and the HRRP anticipation periods, and the HRRP anticipation and HRRP penalty periods (**Supplementary Appendix**).

### Simulation analyses

We tested the hypothesis that comparing average mortality rates would not be able to correctly identify the timing of a change in mortality. Our hypothesis was based on an algebraic proof indicating that if within-period temporal trends exist, ignoring such temporal trends may falsely identify between-period differences associated with the intervention (**Supplementary Appendix**). We created data simulations for mortality, wherein we created a data set in which mortality increased linearly by 1% over the study period, consistent with the overall increase in HF mortality over this period (increase by 0.008% per month starting at 7% in the first month). We explicitly accounted for random variation in monthly rates.^18^ In a positive control, we introduced an increase in mortality such that slope was 50% higher rate after HRRP announcement in April 2010 compared with the slope before the announcement, i.e. slope increased to 0.012% per month (**Figure S1**). We created two negative controls wherein the mortality slope changed 12 months and 24 months before HRRP announcement, in April 2009 and April 2008, respectively (**Figures S2 and S3**). We assessed whether the strategies accurately identified the association between HRRP announcement and mortality trends in the 3 scenarios above.

The analyses were performed using SAS, version 9.4 (Cary, NC), and Stata 16 (College Station, TX). The level of significance was set at 0.05. The codes for the simulation analyses are posted on GitHub.^18^ The study was approved by the Yale University Institutional Review Board.

## RESULTS

We identified 4, 313,523 hospitalizations for HF, 1,788,219 for AMI during the study period, and 3,758,111 for pneumonia. In April 2005, there were 49,043 for HF, 18,705 for AMI, and 50,622 for pneumonia, with a decrease in the number of hospitalizations for all 3 conditions through March 2015.

### Experiment 1: Mortality patterns in period-averaged vs monthly assessments

Across the 4 study periods, unadjusted post-discharge 30-day mortality for both HF and pneumonia increased, whereas unadjusted mortality for AMI decreased in the HRRP anticipation and penalty periods. In unadjusted analyses, after ascribing changes in mortality rates between the baseline and pre-HRRP periods to secular mortality trends, there was no significant excess change between the pre-HRRP and HRRP anticipation periods or the HRRP anticipation and HRRP penalty periods for HF or pneumonia, whereas mortality for AMI decreased (**Figure 2, Tables S1-S3**). In the second strategy, monthly rates examined in interrupted time series models for changes across periods did not find any inflections in unadjusted mortality rates during the HRRP anticipation and penalty periods (**Figure 3, Tables S4-S6**). However, for risk-adjusted rates, period-wise analysis and monthly results produced different results. The period-wise analysis found a significant increase in mortality between the pre-HRRP and HRRP anticipation periods for both HF and pneumonia, without significant changes in mortality for AMI. In contrast, the monthly modeled slope-change interrupted time series models revealed no inflection in mortality slopes for either HF or pneumonia at HRRP announcement. These contrasting conclusions between period-wise and monthly assessments in mortality rates persisted for each of covariate selection strategies for risk-adjustment, compared head-to-head (**Table 1**). We also identified a within period trend in mortality.

**Table 1:**
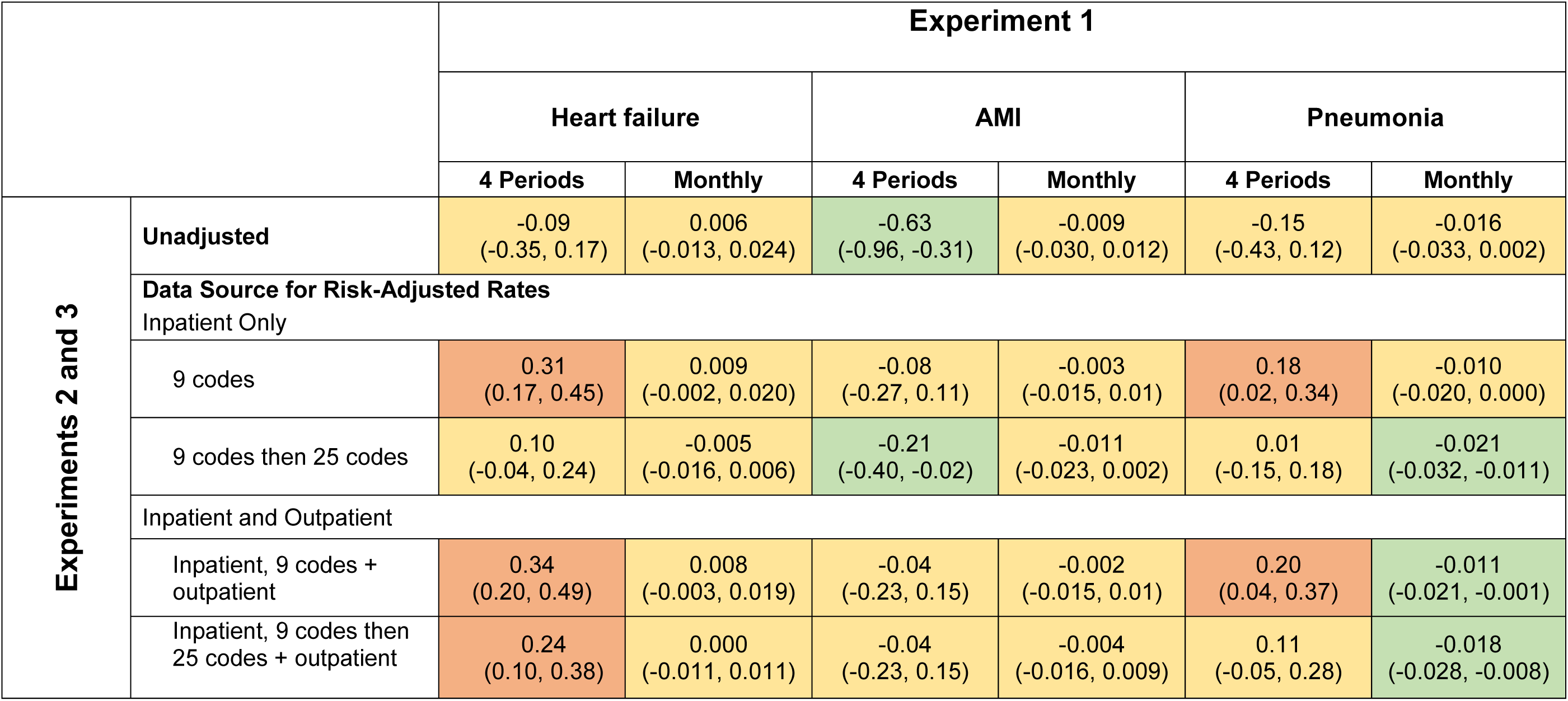
Summary of results of data experiments. Results indicate changes in mortality at the announcement of the Hospital Readmissions Reduction Program. Values under “4 periods” below represent excess changes in consecutive 30-month periods surrounding HRRP announcement. The “monthly” rates represent a change in the slope of the monthly mortality rates at the announcement of HRRP, relative to the slope of mortality in the 30-month preceding HRRP announcement. Note that the confidence intervals are not adjusted for multiple testing. (Colors: Orange: significant increase in mortality, yellow: no significant change, and green: significantly decreased mortality).

**Figure 2:**
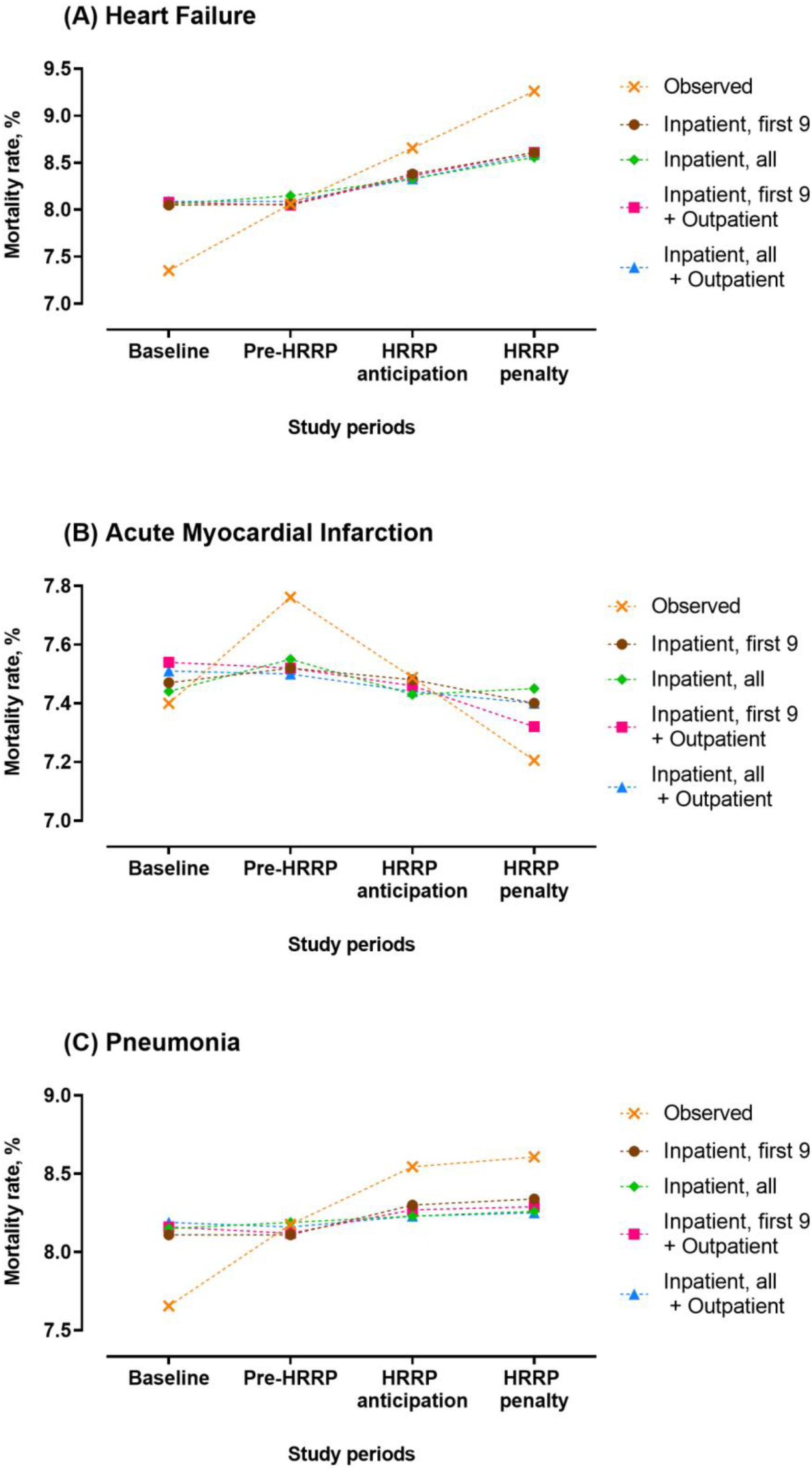
Trends in post-discharge mortality across the 4 study periods, ignoring within-period temporal trends.

**Figure 3:**
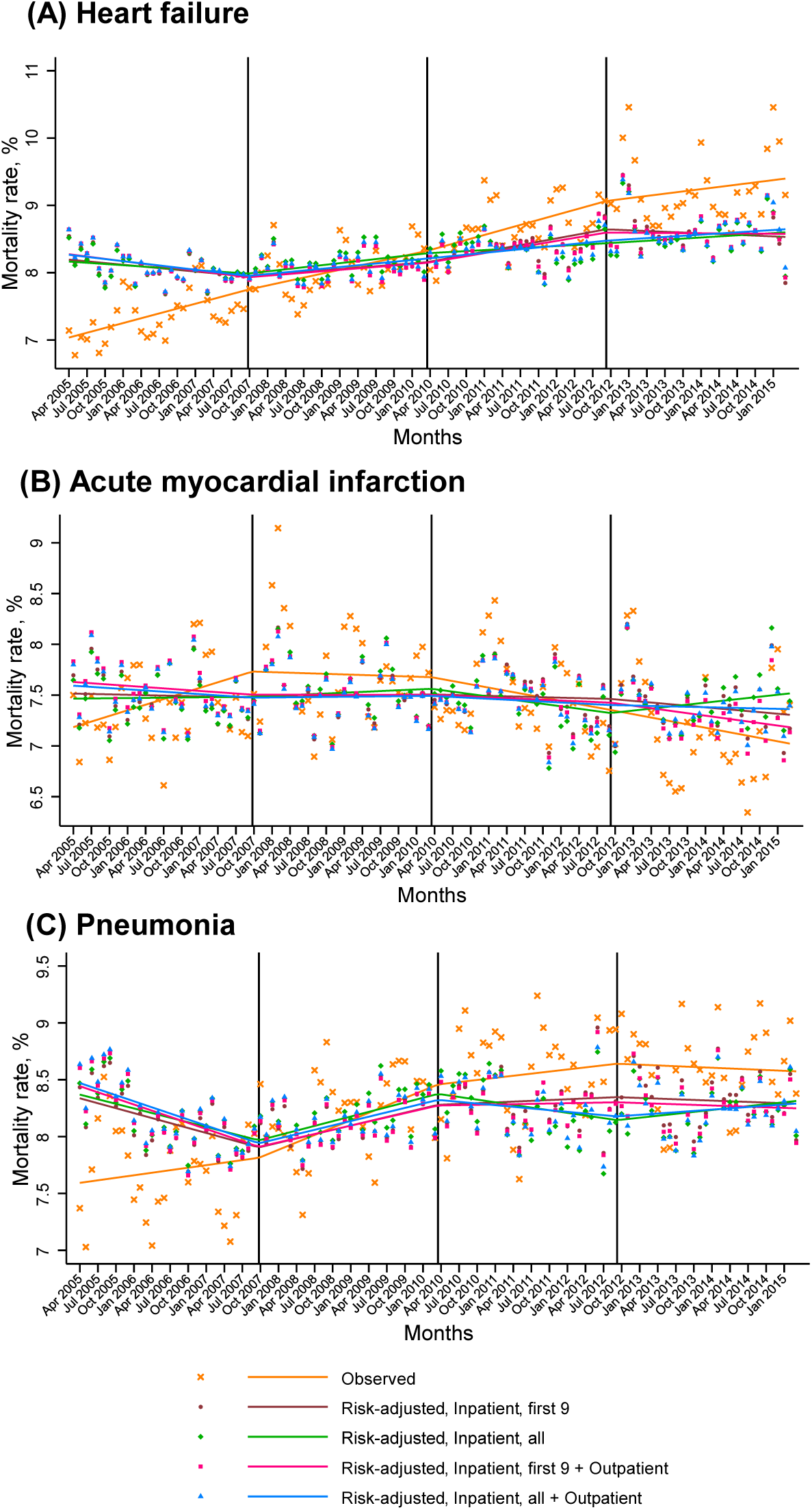
Changes in monthly mortality rates in an interrupted time series framework that accounts for within-period temporal trends. Vertical lines demarcate four 30-month periods: baseline (April 2005-September 2007), pre-HRRP (October 2007-March 2010), HRRP anticipation (April 2010-September 2012), and HRRP penalties (October 2012-March 2015)

### Experiment 2: Risk-adjustment with Inpatient vs Inpatient and Outpatient Claims

The use of outpatient claims identified ∼1.5-fold higher number of risk factors in the baseline period compared with inpatient claims alone for all HRRP conditions (**Figures S4-S6**). There were a mean 9.2, 6.0 and 8.6 risk factors based on inpatient and outpatient claims in HF, AMI and pneumonia, compared with 5.8, 3.8 and 4.8 using inpatient claims alone in the respective groups. For period-wise or monthly strategies there were no differences in the association of HRRP with mortality, regardless of the use of inpatient or inpatient and outpatient data (**Table 1**).

### Experiment 3: Changes in Number of Codes on Inpatient Claims

The number of risk factors remained consistent after code slots increased if only 9 inpatient codes were used, regardless of whether outpatient codes were used. The use of all available inpatient codes, when code slots increased, was associated with an increase in the number of risk factors captured from claims between the pre-HRRP and the HRRP anticipation periods (when the slots increased) but was smaller when outpatient claims were also used (**Figure S4-S6**). The covariate selection strategies, however, did not cause a change in the average mortality risk score across periods or a change in monthly mortality risk score across these periods. There was a steady increase in risk over time in all covariate selection strategies for all conditions, without inflections between pre-HRRP and HRRP anticipation periods, when the number of captured codes increased (**Figure S4-S6**). All covariate selection strategies to identify risk factors for risk-adjustment models produced qualitatively similar results, regardless of whether a fixed number of inpatient claims or a combination of inpatient and outpatient claims were used to adjust for risk over time (**Figure 4, Tables S1-S3**).

**Figure 4:**
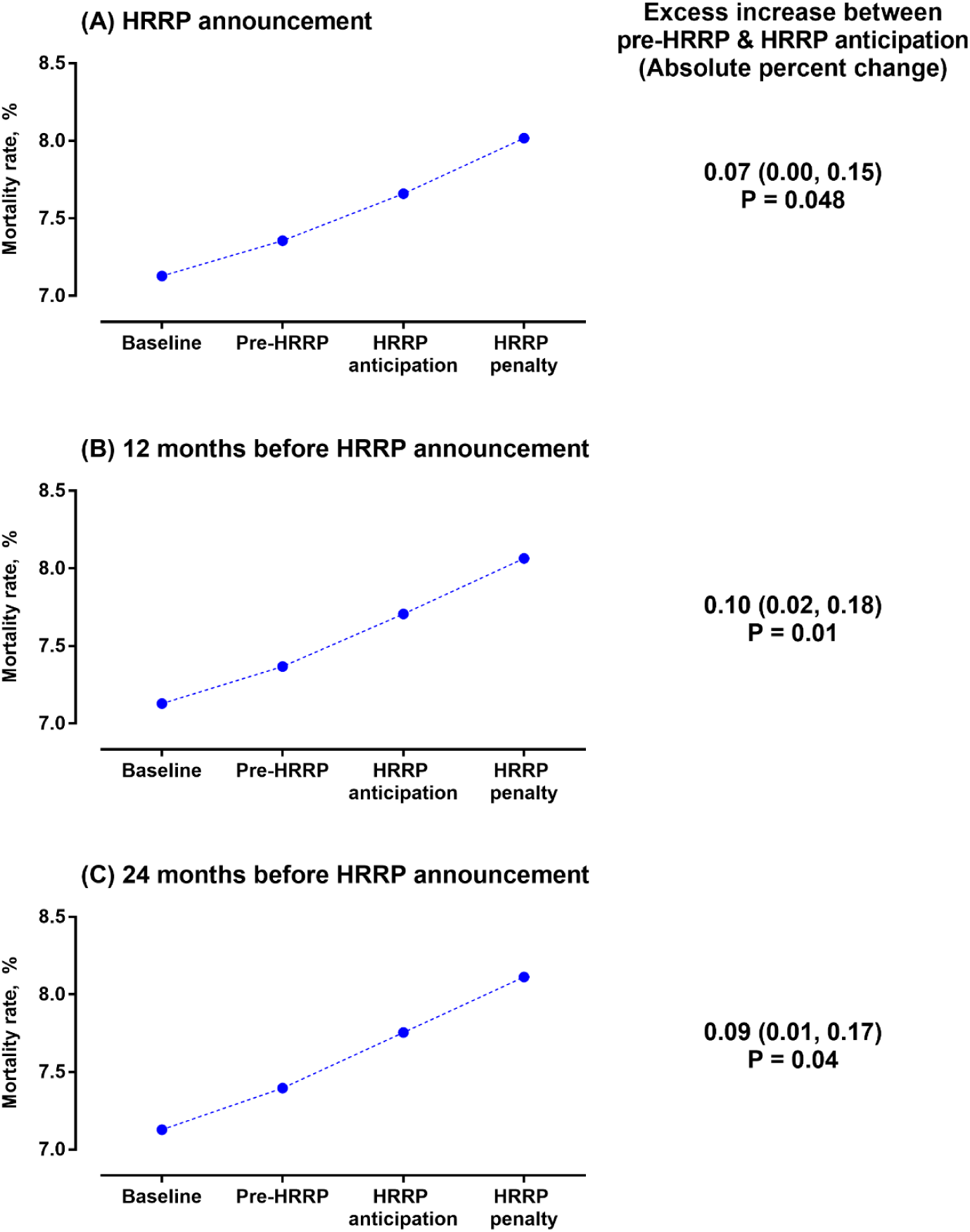
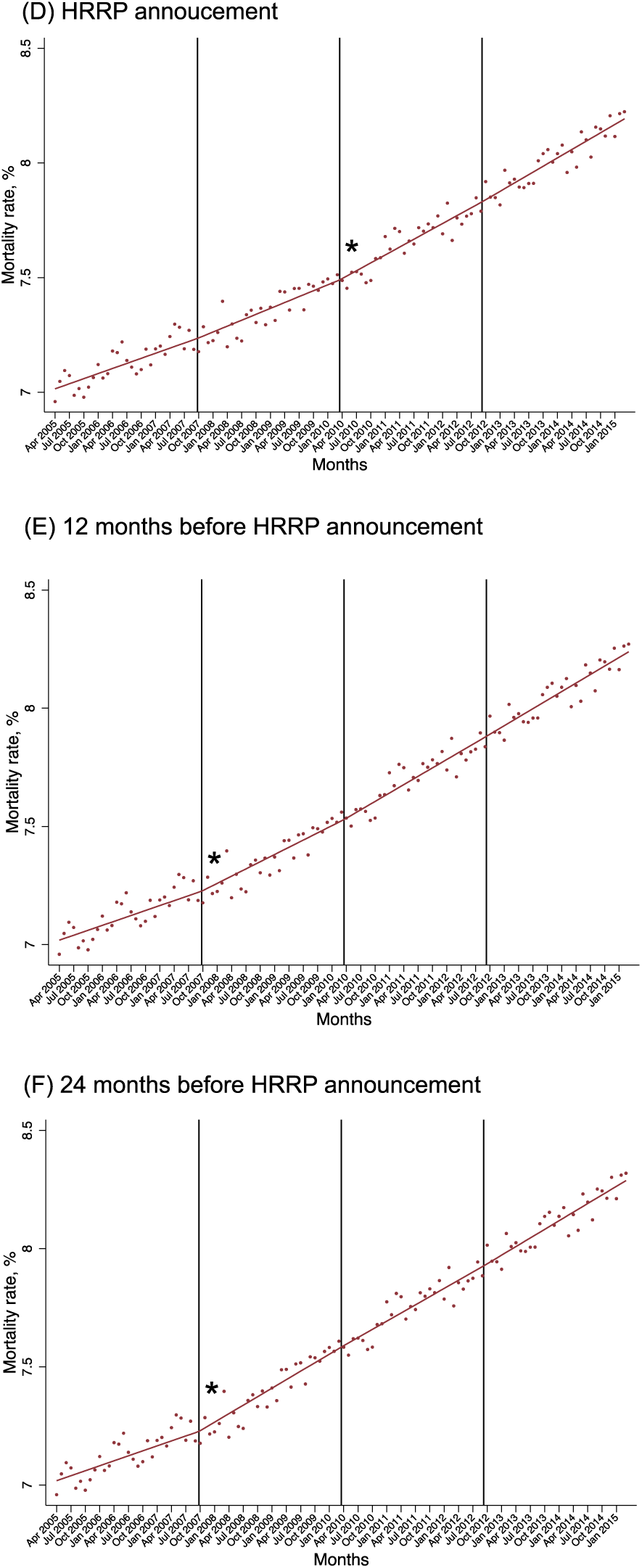
Simulated inflections in mortality rates. **Period-wise mortality assessments (A-C)**. Changes in mortality between the pre-HRRP and the HRRP anticipation periods in excess of the secular changes in mortality between baseline and pre-HRRP periods. Simulated increases in mortality at (A) HRRP announcement in April 2010, (B) 12 months before HRRP announcement in April 2009, and (C) 24 months before HRRP announcement in April 2008.**Monthly rates in interrupted time series models (D-F)**. Changes in slopes of monthly mortality trends between baseline, pre-HRRP, HRRP anticipation and HRRP penalty periods. Simulated increases in mortality at (D) HRRP announcement in April 2010, (E) 12 months before HRRP announcement in April 2009, and (F) 24 months before HRRP announcement in April 2008. *indicates significant changes in slope (P<0.05).

### Simulation analyses

In analyses where we simulated an increase in mortality at HRRP announcement, period-wise and monthly modeled interrupted time series accurately identified that a change occurred at HRRP announcement in April 2010 (**Figure 5**). However, in negative controls with simulated increases preceding HRRP by 12 or 24 months, a period-wise analysis, ascribed the increase in mortality to HRRP announcement (**Figure 5**). In contrast, monthly trends accurately identified that an increase in mortality occurred between the baseline and pre-HRRP periods, without any inflections in mortality at HRRP announcement (**Figure 5, Table S7**).

## DISCUSSION

In our study, we show that in studies evaluating the HRRP with CMS the use of period-specific temporal trends found no association of the HRRP with mortality. However, studying the effect of HRRP as changes in average risk-adjusted mortality rates suggested an association with increased mortality. Furthermore, we identified a problem in the latter method, explaining the discordance in results. The violation of a central assumption led to a misspecification of the timing of the change in mortality, which we found algebraically, and in our analysis using simulations. Different data sources and different strategies to identify covariates for risk adjustment did not further substantially change the finding, particularly for heart failure and AMI.

Our findings validate concerns about the taking average results in four large time periods to make an inference about when changes in rates occurred. In the absence of a control group, such a pre-post health policy intervention evaluation must account for temporal trends in the outcome within the pre and post periods. Evaluating changes in averages if there is no within-period trend is satisfactory. However, ignoring such trends can misattribute the timing of the change. The proof of why this approach can be misleading is best illustrated by algebra (outlined in Online Supplement). The error term in these assessments is directly proportional to the magnitude of trend within periods relative to the trend across periods. In the case of HRRP, mortality was rising in the period before the HRRP was announced, which was also noted previously.^3,19^ Thus, the within-period trend does not satisfy the assumptions of the approach and should preclude its use. This issue has been raised in studies of methodology for more than 30 years.^13^

This issue has contemporary relevance because the study based on averages within the four large time periods has sparked calls for rescinding the policy.^20^ In an oped, the authors suggested that the HRRP resulted in the deaths of 10 thousand patients,^21^ and in another for limiting the expansion of the program,^22^when it is set to target more conditions.^23,24^ The White House has called for an investigation of quality measures, in part as a result of the study. Meanwhile, the Medicare Payment Advisory Commission (MedPAC) concluded that HRRP saved over $2 billion and reduced readmissions without an increase in mortality.^1,6^

Other peer-reviewed studies have evaluated HRRP, but they focused on registry data,^8^ or on trends in specific patient groups,^4,9^ and may not be generalizable to the entire Medicare population.^19,25^ Assessments of hospitals that reduced readmissions have not demonstrated any inflections in mortality.^5,10,16^ An evaluation by MedPAC was consistent with the changes in slopes analysis and found no increase in mortality associated with HRRP.^1,6^ They recently reaffirmed their initial analysis with a report in September 2019.^12^

Our study has several limitations. First, we do not replicate the exact approach used by the study that focused on period-wise assessments because we could not access the code for their analyses, and observations, such as HRRP-associated increase in unadjusted HF mortality, could not be replicated. Similarly, while we were able to replicate the association of HRRP announcement and mortality changes in pneumonia, we could not replicate the association with HRRP implementation. Moreover, for pneumonia, even the association with HRRP announcement varied between unadjusted and risk-adjusted assessments, and various covariate selection strategies. We are unable to explain these differences, especially as we replicate the association of HRRP and mortality for both heart failure and AMI accurately. Second, we do not focus on alternative ways to risk-adjust, such as the inverse probability weighting that were used in the study that suggested HRRP’s association with mortality. We did this intentionally as the study using this approach reported consistent results with logistic regression.^5^ Moreover, the inverse probability weighting model used in the study is itself prone to errors, especially as currently described in the study.^5^Specifically, the authors describe multiple different logistic regression models for comparison between different periods, rather than a common model that included all 4 periods. The former approach would, by design, generate different coefficients for different across-period comparisons, thereby compromising risk-adjustment. Finally, we could not elucidate whether temporal changes in coding practices underlie the observed changes in coded severity of illness. However, risk-adjusted analyses were largely consistent regardless of the number of codes used and did not affect the interpretation of temporal trends, especially for heart failure and AMI. In a separate assessment, we have found that the additional codes from inpatient claims after code slot expansion were largely limited to covariates with relatively limited contribution to measured severity of illness.^26^

In conclusion, policy evaluation that evaluated the HRRP by changes in average rates over periods ignored within-period temporal trends and likely found a spurious association. The concern about this approach, when there are intra-period trends, has been documented in the literature and can be shown with algebraic equations and are evident in the Medicare data and simulations. Thus, more appropriate analyses of the changes in slope fail to reveal an increase in mortality associated with the announcement or implementation of the HRRP.

## Data Availability

The data are proprietary of the Centers for Medicare and Medicaid Services (CMS), and can be obtained from them directly under a data use agreement with them.

## ACKNOWLEDGMENT

### Funding

Dr. Khera is supported by the National Center for Advancing Translational Sciences (UL1TR001105) of the National Institutes of Health. The funder had no role in the design and conduct of the study; collection, management, analysis, and interpretation of the data; preparation, review, or approval of the manuscript; and decision to submit the manuscript for publication.

### Disclosures

Drs. Krumholz, Normand, Bernheim, Lin, and Mr. Wang work under contract with the Centers for Medicare & Medicaid Services to develop publicly reported quality measures. Dr. Krumholz was a recipient of a research grant, through Yale, from Medtronic and the U.S. Food and Drug Administration to develop methods for post-market surveillance of medical devices; is a recipient of a research grant with Medtronic and Johnson & Johnson, through Yale, to develop methods of clinical trial data sharing; was a recipient of a research agreement, through Yale, from the Shenzhen Center for Health Information for work to advance intelligent disease prevention and health promotion; collaborates with the National Center for Cardiovascular Diseases in Beijing; received payment from the Arnold & Porter Law Firm for work related to the Sanofi clopidogrel litigation and from the Ben C. Martin Law Firm for work related to the Cook IVC filter litigation; receives payment from the Siegfried & Jensen Law Firm for work related to Vioxx litigation; chairs a Cardiac Scientific Advisory Board for UnitedHealth; is a participant/participant representative of the IBM Watson Health Life Sciences Board; is a member of the Advisory Board for Element Science, the Advisory Board for Facebook, and the Physician Advisory Board for Aetna; and is the founder of HugoHealth, a personal health information platform and a co-founder of Refactor Health, an enterprise healthcare AI-augmented data management company. Dr. Khera reports no potential conflicts of interest.

The study was conceived and conducted by the authors, and the Centers for Medicare & Medicaid Services played no role in its design and conduct; collection, management, analysis, and interpretation of the data; and preparation, review, or approval of the manuscript.

